# COVID-19 pandemic: Analyzing of different pandemic control strategies using saturation models

**DOI:** 10.1101/2021.04.22.21255952

**Authors:** Stefan Bracke, Lars Grams

## Abstract

Since December 2019, the world is confronted with the outbreak of the respiratory disease COVID-19. At the beginning of 2020, the COVID-19 epidemic evolved into a pandemic, which continues to this day. Within many countries, several control strategies or combinations of them, like restrictions (e.g. lockdown actions), medical care (e.g. development of vaccine or medicaments) and medical prevention (e.g. hygiene concept), were established with the goal to control the pandemic. Depending on the chosen control strategy, the COVID-19 spreading behavior slowed down or approximately stopped for a defined time range. This phenomenon is called saturation effect and can be described by saturation models: E.g. a fundamental approach is Verhulst (1838). The model parameter allows the interpretation of the spreading speed (growth) and the saturation effect in a sound way. This paper shows results of a research study of the COVID-19 spreading behavior and saturation effects depending on different pandemic control strategies in different countries and time phases based on Johns Hopkins University data base (2020). The study contains the analyzing of saturation effects related to short time periods, e.g. possible caused by lockdown strategies, geographical influences and medical prevention activities. The research study is focusing on reference countries like Germany, Japan, Denmark, Iceland, Ireland and Israel.

## 1. Introduction

Since December 2019, the world is confronted with the outbreak of the respiratory disease COVID-19. At the beginning of 2020, the COVID-19 epidemic evolved into a pandemic, which continues to this day. The incredible speed of the spread and the consequences of the infection had a worldwide impact on societies, health systems and social life. Within many countries, several control strategies or combinations of them, like restrictions (e.g. lockdown actions), medical care (e.g. development of vaccine or medicaments) and medical prevention (e.g. hygiene concept), were established with the goal to control the pandemic. Depending on the chosen control strategy, the COVID-19 spreading behavior slowed down or approximately stopped for a defined time range. This phenomenon is called saturation effect and can be described by saturation respectively growth models. A fundamental approach is the Verhulst model (1838) which contains the parameter saturation limit among other parameters. The model parameter allows the interpretation of the spreading speed (growth) and the saturation effect in a sound way. A limitation of using these models is the time period in which growth can be well represented. The COVID-19 pandemic phase runs over a long period of time (12.2019 until today (02.2021) and the spreading behavior is changing frequently, e.g. caused by many different activities within the different control strategies or seasonal effects.

This paper shows results of a research study of the COVID-19 spreading behavior depending on different pandemic control strategies in different countries and time phases. The study contains the analyzing of saturation effects related to short time periods, e.g., possibly caused by lockdown strategies, geographical influences and medical prevention activities. The research study contains case studies focusing on the data base of reference countries.

## 2. Goal of Research Study

The overarching goal of the research study is the analysis of the saturation effects with regard to infection occurrence of COVID-19 due to different pandemic control strategies. The detailed goals are as follows:

1. Analyzing of saturation effect with regard to the first, second and third wave (time range: 12.2019 until 02.2021).
2. Comparison of measures of the government regarding pandemic control (to reduce peak/waves), which can be the reason for saturation effects
3. Comparison of spreading behavior (based on normalized time ranges) and saturation effects with regard to different countries: Germany, Japan, Denmark, Iceland, Ireland and Israel.

## 3. Base of operations: Data and information

### 3.1 Data source

The data used for the presented research study is taken from the JHU COVID-19 Dashboard by Dong et al. (2020). (Dong, Du, and Gardner 2020). The COVID-19 Dashboard provides worldwide infection data since the 01-22-2020 for infected, dead and recovered cases.

The cumulated numbers of infected cases of six countries are analyzed to detect multiple infection waves in a time period of 420 days from 01-22-2020 until 03-16-2021 regarding each country. The countries chosen are Germany, Japan, Denmark, Ireland, Iceland and Israel. The different waves were separated by the minimum daily cases between two periods of high daily infection cases respectively two waves. The reversal of the trend at the minimum point can also be substantiated by the application of the Cox and Stuart trend test (Cox and Stuart 1955). Thus, two infection waves were identified for Germany and Iceland, while the course of infection for Denmark, Japan, Ireland and Israel could be distinguished in three infection waves each.

As a basis for comparison, the population, population density and relative number of infected people were considered.

### 3.2 Data uncertainty

It must be considered, that there are uncertainty factors in the data base. Here a brief overview of the uncertainty with regard to data acquisition is given, for a detailed overview cf. Bracke et al. (2020) and Puls and Bracke (2020). First of all, the type of measuring method has to be considered. Three aspects are important:

- Criteria for testing (test strategy, e.g., symptom-based or area-wide),
- Reporting system (reporting procedure),
- Accessibility of health department (e.g., weekend-impact).

Furthermore, apart from the lockdown measures taken, the spreading is also influenced by the dynamic occurrence of infections and by the handling of the pandemic. Some of these uncertainty factors are (without claiming to be conclusive):

- Seasonality and climatic effects, cf. (Sajadi et al. 2020),
- Mutations of the virus,
- Type of treatment, cf. (Gattinoni et al. 2020) and
- Vaccination progress.

### 3.3 Measures taken for selected countries

During the period of the first infection wave around April, all selected countries except Japan and Iceland declared a lockdown, which was then followed by loosenings as the number of daily infections decreased. Other measures taken were travel limitations, face mask obligation, social distancing and cancelling of bigger events.

## 4. Fundamentals: Definitions and Method

This section provides an overview of the used methodology as well as aspects of data uncertainty, which have to be taken into consideration. The logistic function or Verhulst function is used to model the different infection waves. Parameters were estimated using least squares estimation.

### 4.1 Logistic/Verhulst Function

The different infection waves show an exponential increase of infected people in the beginning, before reaching a certain saturation value, this behavior results in the characteristic S-shaped curve, which was first discovered by Verhulst(Verhulst 1938) to describe the growth of populations and can be expressed by Eq. (1).

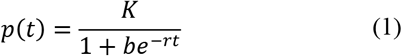

Here K describes the saturation limit, where the number of infections is striving against. The factor r describes the growth rate, while b impacts the starting point and therefore the translation on the x-axis, since Eq. (2) is fulfilled.

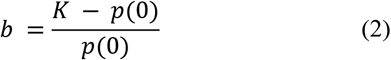

In Figure 1 a logistic function with the characteristic S-shape is shown.

**Fig. 1.**
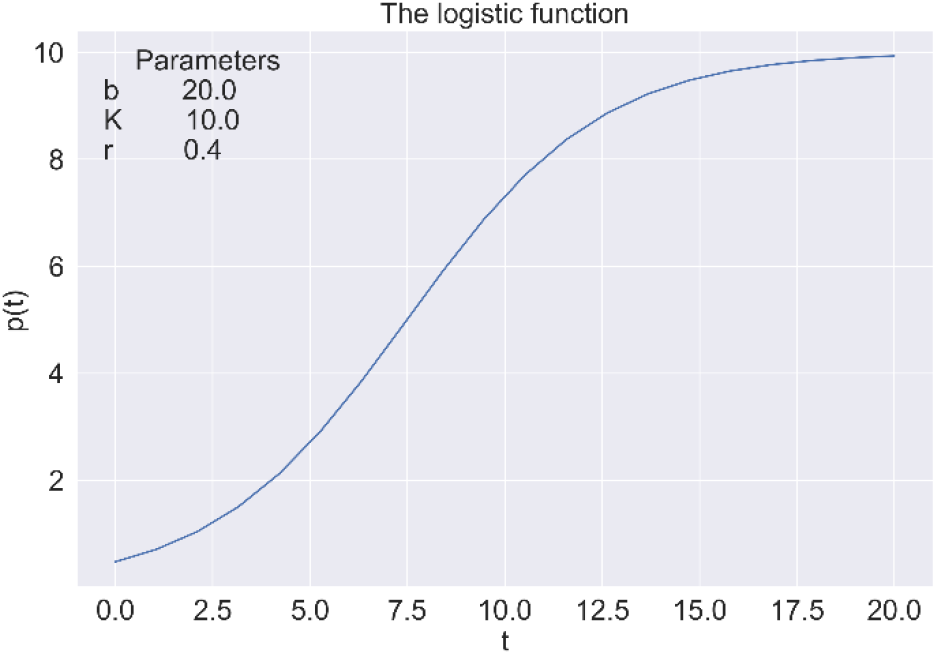
The logistic function with synthetic parameters.

The logistic function in Figure 1 has a growth rate r = 0.4 and saturation limit K = 10. The b parameter yields a value of b = 20.

## 5. Data Analytics

The following section includes the course of infection including the days, where different waves were separated (red vertical lines). Saturation models based on the logistic function based on Eq. (1) were used.

### 5.1. Saturation models

The different waves for each country are normalized with respect to the time and number of infected people since the beginning of each wave. By doing this, it is possible to compare different waves with each other. The parameters of the estimated functions for each country are summarized in Table 1 in section 5.2.

**Table 1.** Data base and characteristics of selected countries, time period 01-22-2020 until 03-16-2021, JHU (2020).

| Country | Population [million] | Population Density [population per km<sup>2</sup>] | Infected Cases per 100.000 Inhabitants |
| --- | --- | --- | --- |
| Germany | 83.02 | 233 | 3135.47 |
| Japan | 126.2 | 347.07 | 355.90 |
| Denmark | 5.78 | 137.07 | 11.44 |
| Iceland | 0.357 | 3.5 | 1705.04 |
| Ireland | 4.95 | 70.45 | 4599.25 |
| Israel | 8.88 | 410.53 | 9264.67 |

#### 5.1.1 Saturation models – Germany

The infection course of Germany is divided into two waves.

In Figure 2 it can be seen that after 172 days a second wave starts with an increased number of infected people.

**Fig. 2.**
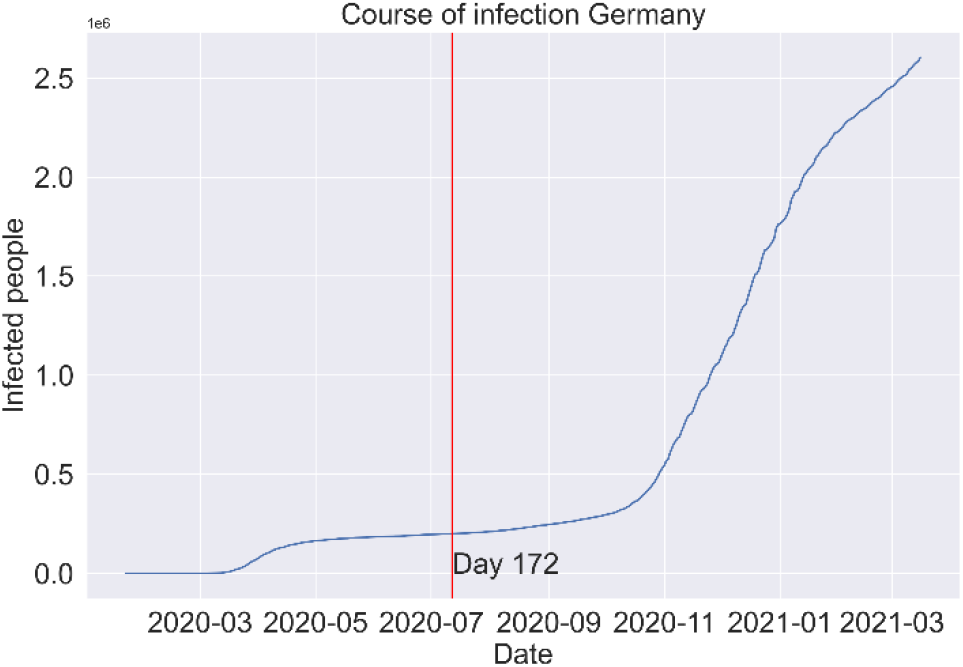
Course of Infection Germany

Figure 3 shows, that the second wave has a much higher impact than the first wave. The number of infected people in the second wave is still increasing. It can also be seen that the saturation model underestimates the number of infected people at the end of the second wave.

**Fig. 3.**
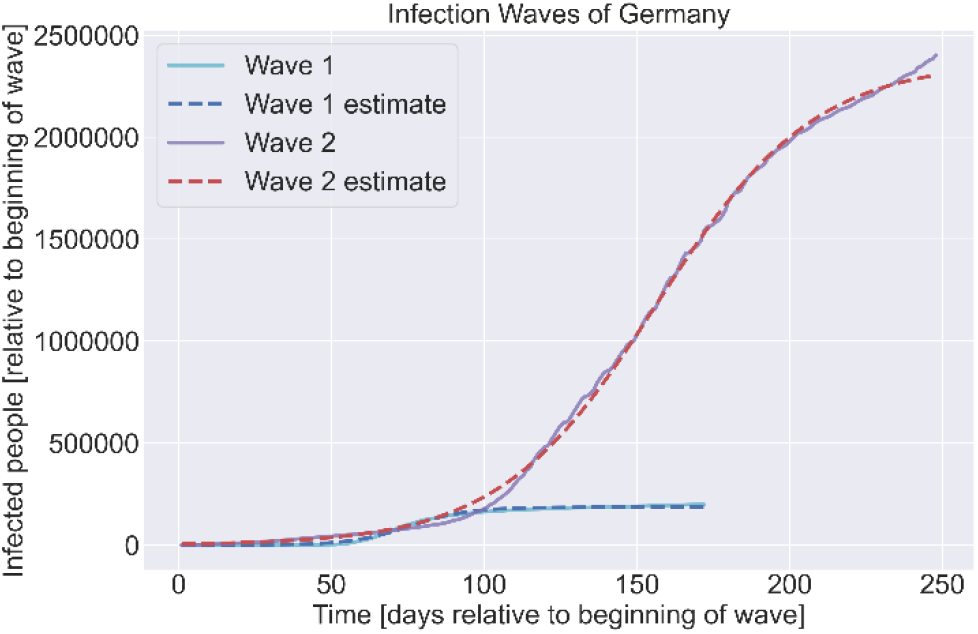
Different infection waves Germany

**Fig. 4.**
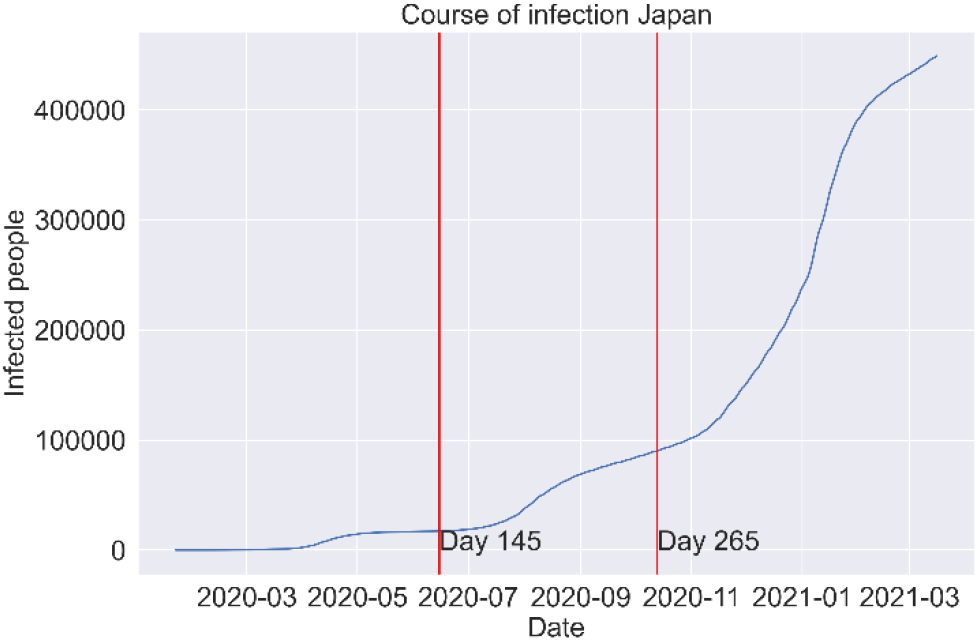
Course of infection Japan

#### 5.1.2 Saturation models – Japan

Japans course of infection can be divided into three waves.

At day 145 and day 265 the start of a new wave can be noticed in Japan.

In Figure 5 it can be seen, that the impact of the waves increased with each wave. It is also worth noting, that the first wave lasts longer than the second wave, but less than the third wave.

**Fig. 5.**
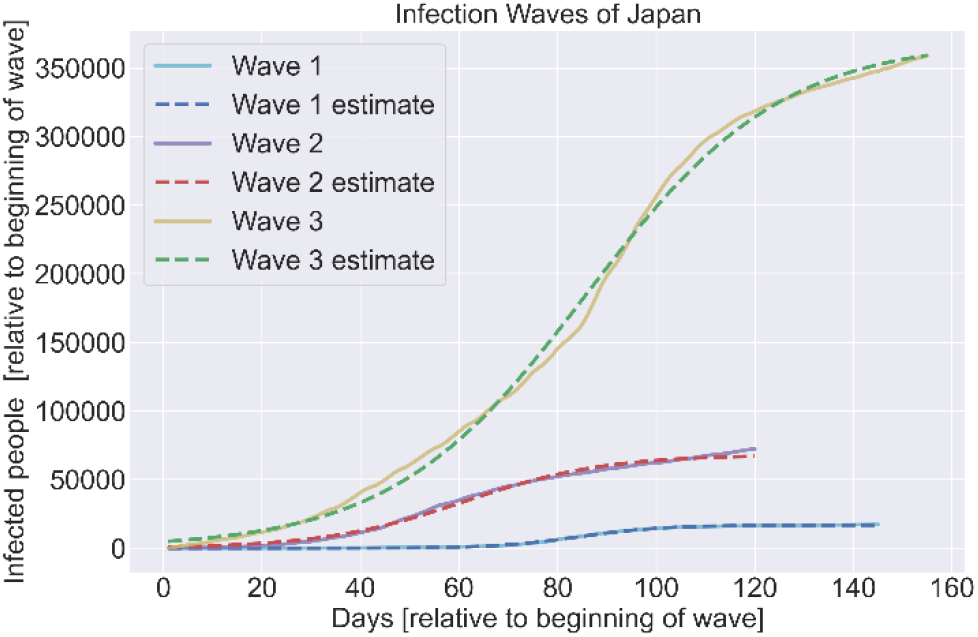
Different infection waves Japan

#### 5.1.3 Saturation models – Denmark

Even though Denmark has very few infected cases, the course of infections can be separated into three waves.

After a long period of almost no infections, a second wave starts at day 130 until day 305 where a third wave can be observed.

The impact of waves is quite different compared to Japan or Germany. The third ongoing wave has the lowest impact, while the second wave has the highest. It is also noticeable that the duration of waves increases when the impact is higher. Furthermore, it stands out, that the plateau between the waves is approximately on a horizontal level, cf. Fig 6. This effect can be a hint for the high effectiveness of the taken measures regarding the pandemic control, because the growth is close to zero.

**Fig. 6.**
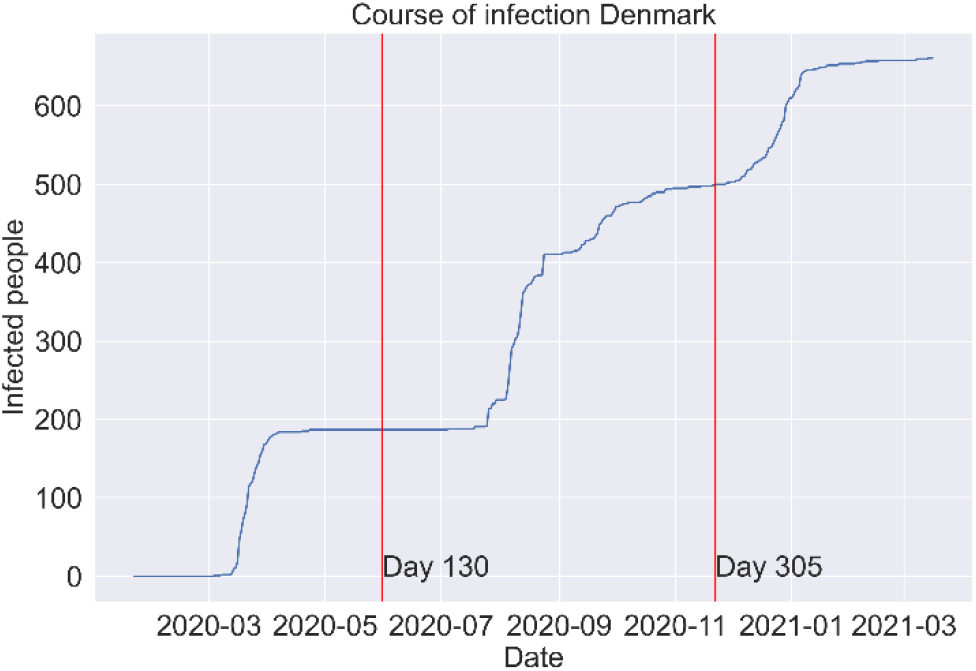
Course of infection Denmark

**Fig. 7.**
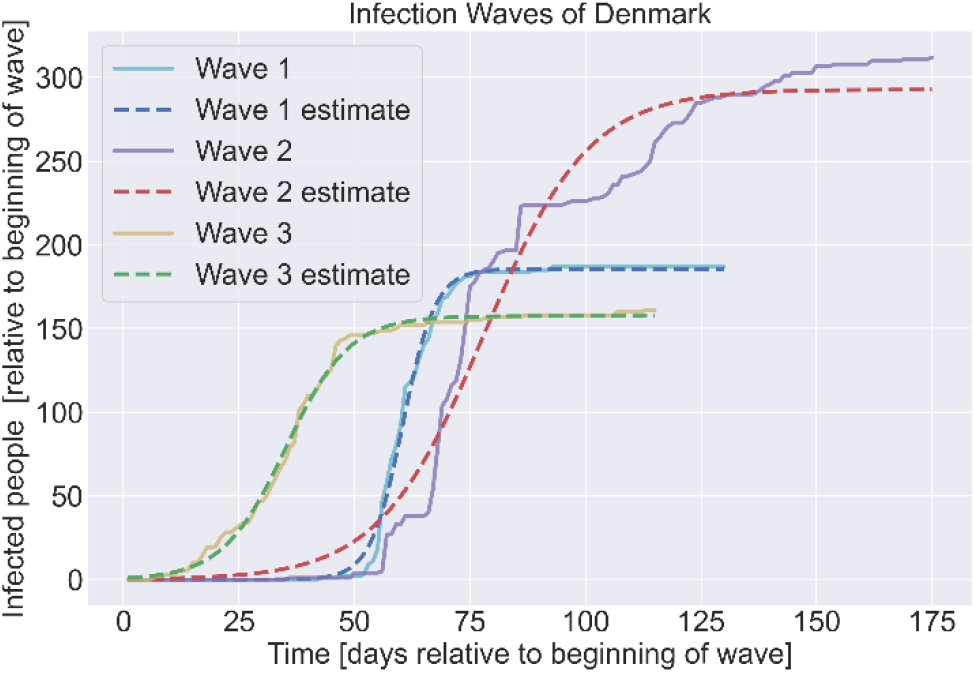
Different Infection Waves Denmark

**Fig. 8.**
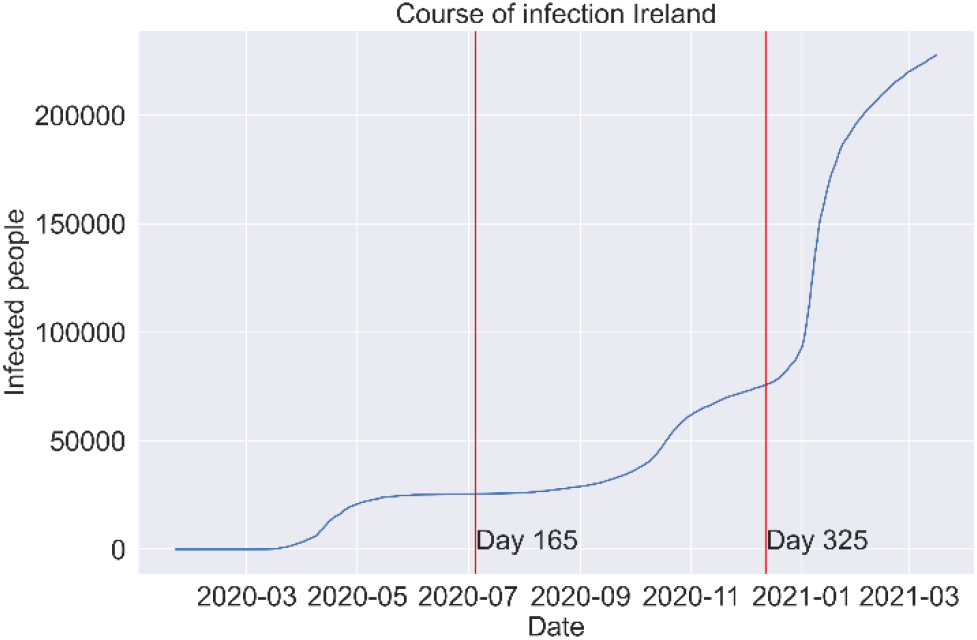
Course of infection Ireland

#### 5.1.4 Saturation models – Ireland

The infection course of Ireland can be separated into three different waves.

The second wave in Ireland starts at day 165 and lasts for almost the same time until at day 325 a third wave with greatly increased number of infected people can be observed.

Figure 9 shows, that even though wave three lasts for less days than wave one and two, the number of infected people in wave three is almost twice as high as in wave one and two together.

**Fig. 9.**
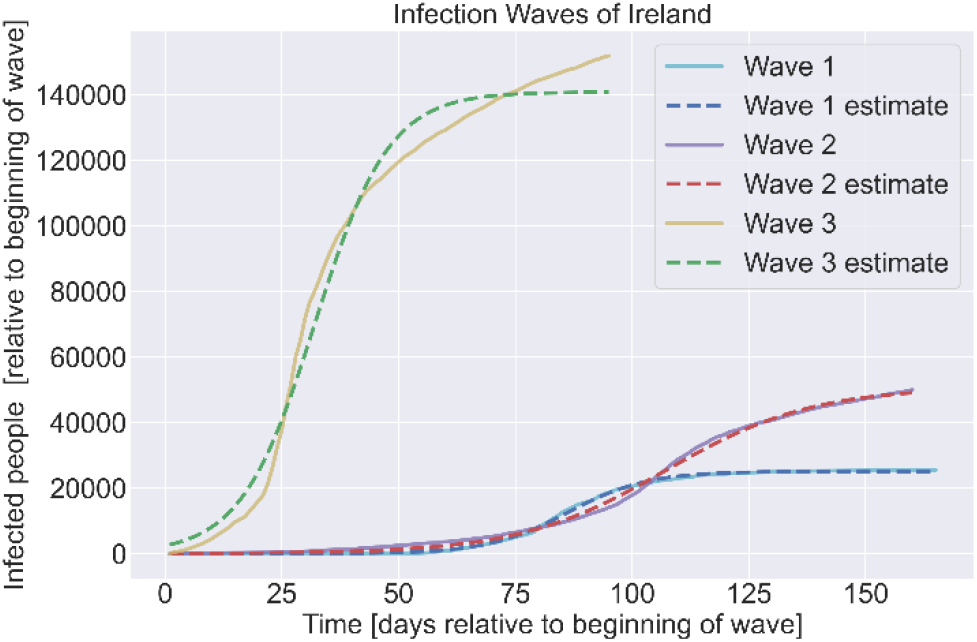
Different infection waves Ireland

#### 5.1.5 Saturation models – Iceland

In Iceland two infection waves can be observed, like in Germany.

In Figure 10 it can be seen, that the second wave starts at day 185.

**Fig. 10.**
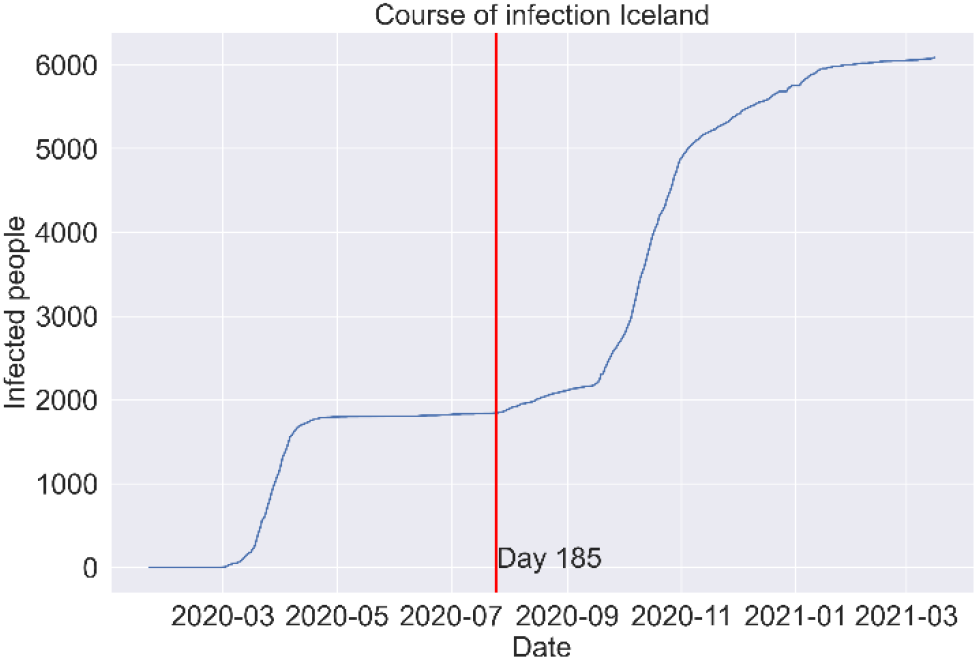
Course of infection Iceland

Figure 11 shows, that in Iceland the second waves lasts longer than the first wave, furthermore more people have been infected. Furthermore, it stands out, that the plateau between the waves is approximately on a horizontal level (this effect is also visible regarding Denmark); cf. Fig. 10. This effect can be a hint for the high effectiveness of the taken measures regarding the pandemic control, because the growth is close to zero.

**Fig. 11.**
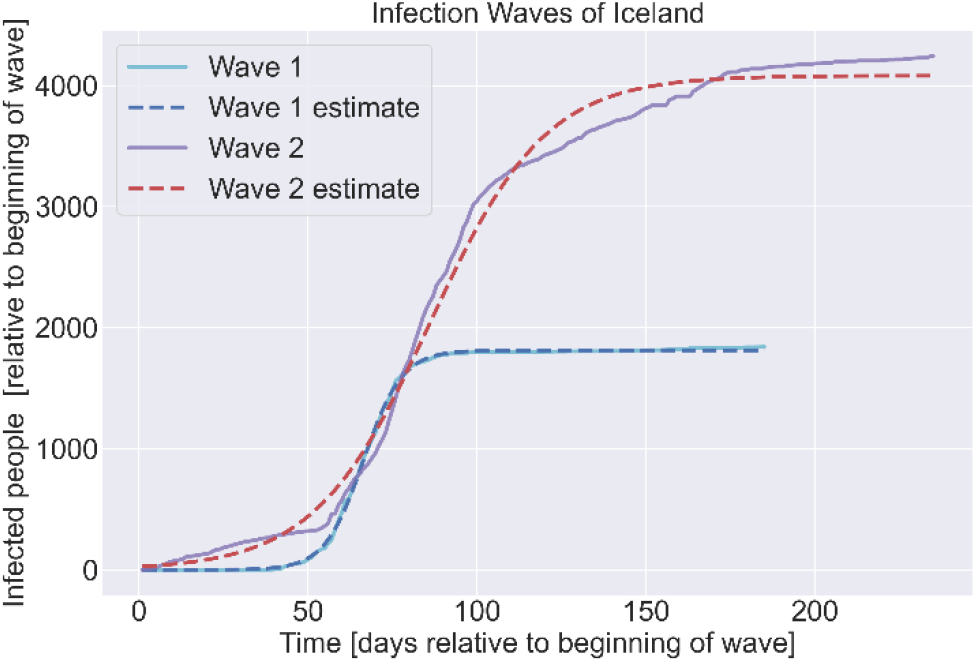
Different infection waves Iceland

**Fig. 12.**
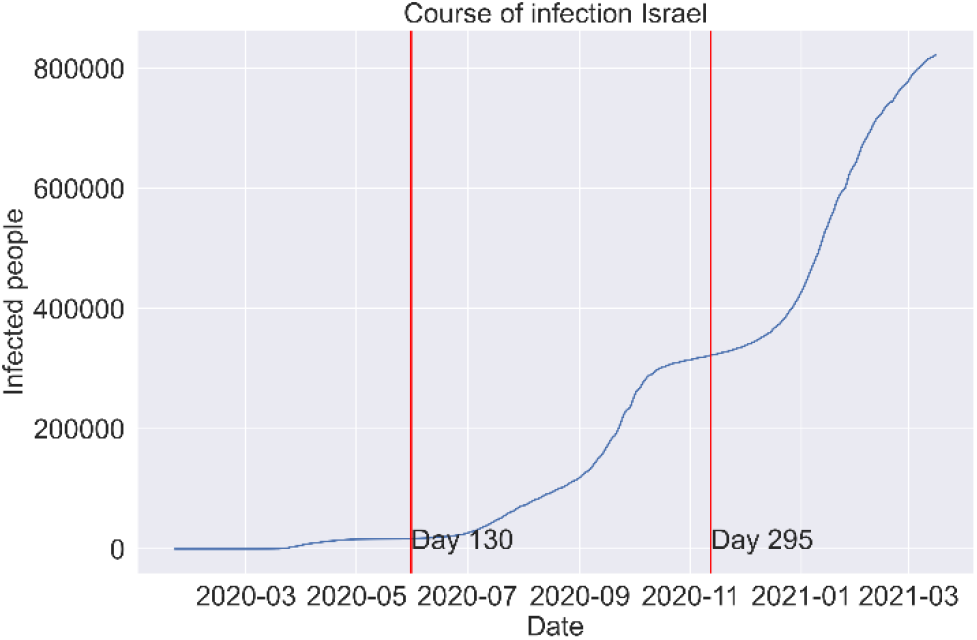
Course of infection Israel

#### 5.1.6 Saturation models – Israel

The infection course of Israel shows three different infection waves.

The days, at which a new wave starts in Israel are similar to the days in Denmark. In Israel the second wave starts at day 130 and the third wave starts at day 295.

Figure 13 shows, that the number of infected people increases with each ongoing wave. It can be observed, that wave two lasts the longest.

**Fig. 13.**
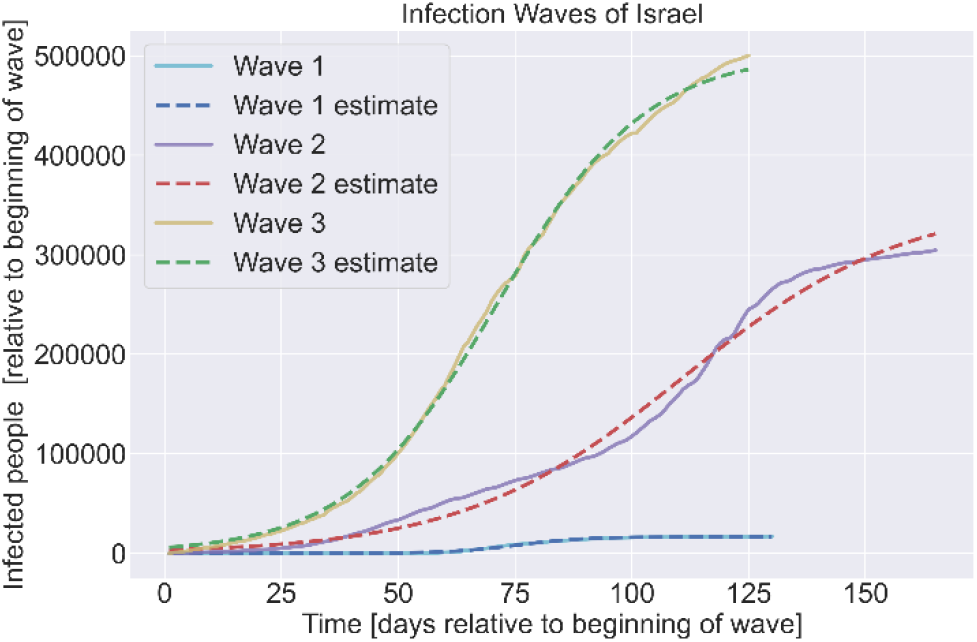
Different infection waves Israel

### 5.2. Comparison of COVID-19 waves via saturation models

Within table 2, the parameters of the estimated logistic functions per analyzed wave and country are listed.

**Table 2.** List of estimated Saturation model parameters with regard to COVID-19 waves related to counties

| Country |  | Wave 1 | Wave 2 | Wave 3 |
| --- | --- | --- | --- | --- |
| Germany | b | 2714.501 | 438.0825 |  |
|  | K | 185831.4 | 2371334 |  |
|  | r | 0.104363 | 0.038825 |  |
| Japan | b | 21764.34 | 72.95367 | 76.27213 |
|  | K | 16797.44 | 68073.36 | 370172.4 |
|  | r | 0.118249 | 0.070312 | 0.050504 |
| Denmark | b | 31478279 | 948.2616 | 169.4984 |
|  | K | 185.7242 | 292.8782 | 157.5402 |
|  | r | 0.285487 | 0.087845 | 0.146056 |
| Iceland | b | 111781.1 | 153.5443 |  |
|  | K | 1813.281 | 4082.059 |  |
|  | r | 0.174984 | 0.058449 |  |
| Ireland | b | 23608.31 | 909.145 | 57.82478 |
|  | K | 25102.4 | 50796.23 | 140841.9 |
|  | r | 0.116753 | 0.063527 | 0.126209 |
| Israel | b | 44522.04209 | 107.3004 | 88.65867 |
|  | K | 16479.9503 | 354105.1 | 502966.8 |
|  | r | 0.141733146 | 0.042105 | 0.062918 |

In Table 2 it can be seen, that the infection rate r of the second wave decreased for all countries and increased again within the third wave. Only Japan being an exception with a constant decreasing infection rate. The increased r-value for the third waves might also occur due to the estimation method, because the estimation leads to a saturation limit close to the last data point given. That way the increasing number of infections is artificially cut off, which means the saturation limit is reached in a shorter time span, which results in a higher r-value.

Figure 14 shows the number of infected people of each wave normalized for each country. Japan, Ireland and Israel all had quite few infections in their first two waves, compared to the number of infections in their third wave. Note that the last wave of each country most likely has not reached a saturation limit yet, therefore the number of infections for that wave is still increasing.

**Fig. 14.**
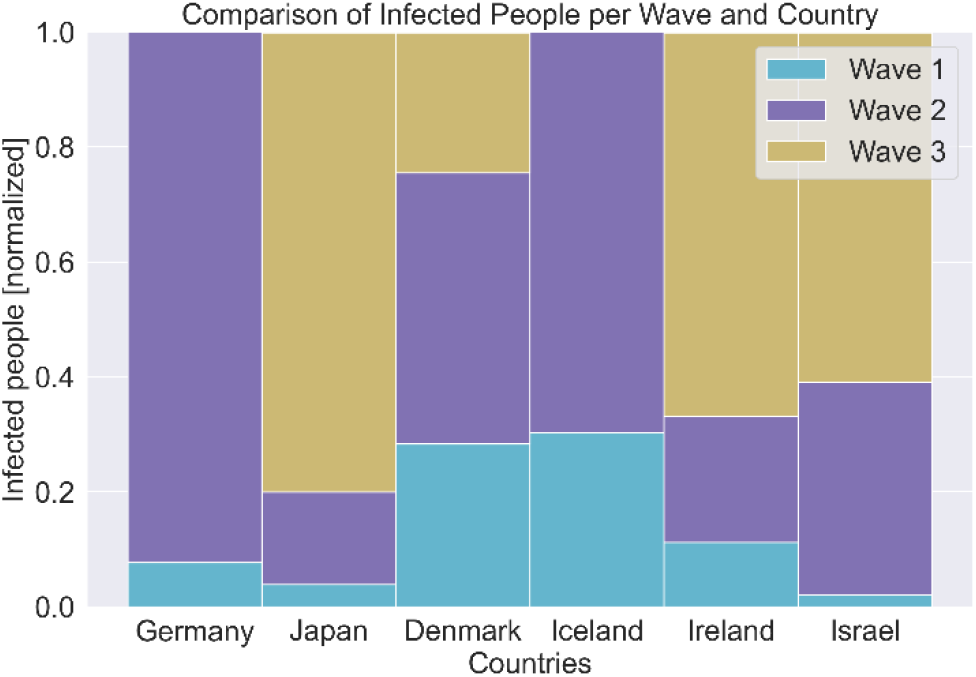
Comparison of infected people per wave and country

Figure 15 gives an overview about the durations of the infection waves for each country. Note that the last wave of each country might still be active. It can be seen that the first wave in Iceland and Germany lasts longer than the first wave in the countries with 3 total infection waves. The duration of the second wave is almost the same for Denmark, Ireland and Israel. The second wave in Japan on the other hand lasted only for around 120 days, which is the shortest duration and can be a hint regarding the effectiveness of the measures to control the pandemic. Therefore, Japan is the first country with a third wave being present.

**Fig. 15.**
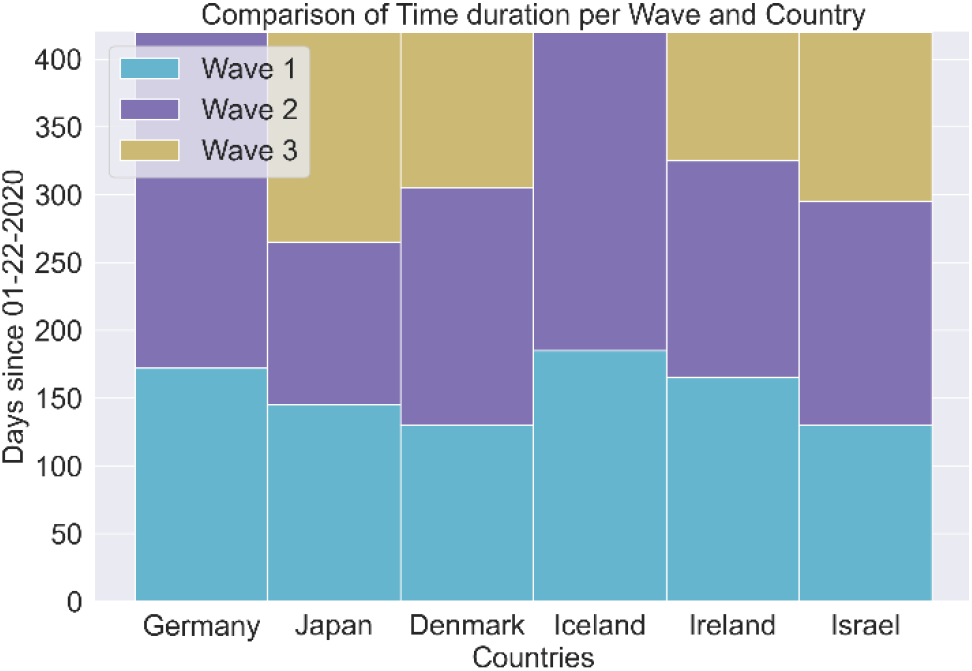
Comparison of time duration per wave and country.

Table 2, Figure 14 and Figure 15 show that the absolute number of infections per wave is increasing even though later waves not only have a lower growth rate r, but also last for almost equal or less the number of days than the previous wave. This can be explained by the increased number of infected persons of the previous wave, those were able to infect more people, that way the absolute number of infected people increased, even though the growth rate r decreased.

## 6. Summary

The COVID-19 pandemic has a huge impact on the whole world. Countries fight the spread of infection with measures like face masks, lockdowns and social distancing. In this paper six countries for which the course of infections showed multiple infection waves where analyzed. It was shown, that the growth r of the second waves, which describes the spreading speed of the infection, decreased in every country. Statements about the growth rate of the third waves yield an error, because the estimation method might estimate the saturation limit too early, which leads to a higher growth rate. Even though the absolute infection numbers increase faster than before, a negative trend regarding the infection speed can be observed from the first to second wave in every country. Furthermore it can be observed, that the plateau between the waves especially in Denmark and Iceland (Fig. 6 and 11) is approximately on a horizontal level. This effect can be a hint for the high effectiveness of the taken measures regarding the pandemic control: The growth stops for a certain time range. This effect is not distinct on the same level in other countries (e.g. Germany, Fig. 2). It can be a hint, that countries in an island position or in comparable position have the advantage to slow down the introduction of the virus from outside based on strict border controls respectively a halt to international (air) traffic. Finally, the length of the waves can be a hint regarding the effectiveness of the measures to control the pandemic, e.g. the second wave in Japan lasted only for around 120 days, which is the shortest duration in comparison to the other analyzed countries.

## Data Availability

all data are from johns hopkins university (jhu) dashboard COVID-19

https://gisanddata.maps.arcgis.com/apps/opsdashboard/index.html#/bda7594740fd40299423467b48e9ecf6

## References

Cox, D. R., and A. Stuart. 1955. “Some Quick Sign Tests For Trend In Location And Dispersion.” Biometrika Vol. 42 (Issue 1-2): 80–95.

Dong, Ensheng, Hongru Du, and Lauren Gardner. 2020. “An Interactive Web-Based Dashboard to Track COVID-19 in Real Time.” The Lancet Infectious Diseases 20 (5): 533–34. https://doi.org/10.1016/S1473-3099(20)30120-1.

Gattinoni, Luciano, Davide Chiumello, Pietro Caironi, Mattia Busana, Federica Romitti, Luca Brazzi, and Luigi Camporota. 2020. “COVID-19 Pneumonia: Different Respiratory Treatments for Different Phenotypes?” Intensive Care Medicine 46 (6): 1099–1102. https://doi.org/10.1007/s00134-020-06033-2.

Sajadi, Mohammad M, Parham Habibzadeh, Augustin Vintzileos, Shervin Shokouhi, Fernando Miralles-Wilhelm, and Anthony Amoroso. 2020. “Temperature, Humidity, and Latitude Analysis to Estimate Potential Spread and Seasonality of Coronavirus Disease 2019 (COVID-19).” Infectious Diseases, 11.

Verhulst, Pierre-Francois. 1938. “Notice Sur La Loi Que La Population Poursuit Dans Son Accroissement.” Correspondance Mathé-Matique et Physique 10: 113–21.

Puls, A. and Bracke, S. (2020). COVID-19 pandemic risk analytics: Data mining with reliability engineering methods for analyzing spreading behavior and comparison with infectious diseases. Proceedings: The International Workshop on Reliability Engineering and Computational Intelligence, Ed. Zaitseva, E. et al, Springer publisher, - in press -

Bracke, S., Puls, A. and Grams, L. (2020). COVID-19 pandemic data analytics: Data heterogeneity, spreading behavior, and lockdown impact. Proceedings of the 30th European Safety and Reliability Conference and the 15 the Probabilistic Safety Assessment and Management Conference. Research Publishing.

